# Open-source C++ Framework of Nonnegative Matrix Factorization and its applications in neuroimaging

**DOI:** 10.1101/2025.11.27.25340327

**Authors:** Youngha Hwang

## Abstract

Non-negative matrix factorization (NMF) produces a factorization that constrains the elements of both the factor matrices to be non-negative. It has been a popular feature extraction method in many applications including neuroimaging. One limitation of the existing softwares of NMF is that they were written in and dependent on the proprietary software of MATLAB. To address this limitation, we introduced an open-source C++ package for performing NMF. To make NMF more accessible to the scientific research community, we describe a NMF algorithm implemented using the Insight Toolkit ITK and Armadillo, a MATLAB style C++ based math library. Armadillo facilitates the computations in linear algebra by calling functions without any need to implement functions in C++. In addition, This framework supports the read and write interface to images specific to neuroscience. Finally, The package supports NMF with multiplicative update and sophisticated initialization methods. We showed that the package has accuracy matching MATLAB and its speed close to that of MATLAB. We used simple simulated images to test its functionality. Then, we demonstrated how the package can be used to analyze neuroimaging data. Specifically, we used the package to find a data-driven set of structural patterns(factor matrices) that are similar across individuals. We validated this factorization method by associating their weighted loading matrices with body mass indices (BMI) of individuals from the human connectome project.

## 1 Introduction

Non-negative factorization (NMF) has been a popular choice of feature extraction due to its interpretability and enforcing sparsity[6, 9]. In neuroimaging area, [16] discussed NMF to analyze structural neuroimaging data. [17] discussed association between grey matter and disease with aging. [7] applied NMF to resting-state functional magnetic resonance imaging (fMRI) to find a functional brain network.

The non-negativity constraints differentiate NMF with respect to the rest of factorization methods and lead to the following two properties of significant interest in medical imaging[16]. First, the non-negativity of the entries of the components matrix matches the expectation of the physical properties of the measured signal. Second, the non-negative entries result in a purely additive combination of the non-negative components. In other words, the data is approximated as a parts-based representation.

There are many publicly available NMF implementations in MATLAB such as [8], NMF Library (https://github.com/hiroyuki-kasai/NMFLibrary), and NMF Toolbox (https://github.com/colinvaz/nmf-toolbox) in addition to the ours (https://github.com/asotiras/brainparts). Even a proprietary function of NMF has been available in MATLAB version 2020a (https://www.mathworks.com/help/stats/nnmf.html). Other implementations include the scikit-learn library from Python[11] and one from the Insight Toolkit (ITK)[14].

However, problems exist of the previous implementations. Firstly, they tend to depend on the proprietary software of MATLAB. So the users must have access to MATLAB before they can use the implementations. Secondly, NMF Toolbox, the implementation in scikit-learn library and the proprietary implementation by MATLAB does not have sophisticated initialization methods such as Non-Negative Double Singular Value Decomposition (NNDSVD) and its variations[1]. Thirdly, C++ based ITK is not easy to use for general public and it is not flexible because it was specially designed for medical image filtering, segmentation, registration, etc. A NMF method implementation in ITK has a limit its expansion in biomedical image processing because different applications require C++ implementation of different metrics, different methods of updating the decomposition matrices, convergence criteria, etc. Finally, the previous softwares do not have any neuroimaing oriented interfaces to Neuroimaging Informatics Technology Initiative (NIFTI) and Connectivity Informatics Technology Initiative (CIFTI), respectively (https://nifti.nimh.nih.gov/, https://www.nitrc.org/projects/cifti/).

Aiming towards a good balance between speed and ease of use, a high quality linear algebra library, Armadillo, has been developed[13]. It is the open-source C++ template wrapper of linear algebra libraries. It provides high-level syntax and functionality deliberately similar to Matlab as shown in the Appendix. Thus, it is useful for quick conversion of research code into production environments. From the ITK-based class framework defining the common interface of NMF[10], an Armadillo-based implementation of a specific method was provided in this article using the Frobenius norm as the underlying objective function. In addition, NIFTI read and write functions were provided from ITK. Similarly, CIFTI read and write functions given from Cifti Library (https://github.com/Washington-University/CiftiLib).

We demonstrated its effectiveness in accuracy and speed against MATLAB along with its versatileness in adopting other metrics, initialization methods, etc. Then, we showed three working examples. One is a self contained unit test showing the method usage. The second test uses a simulated data set corrupted by noise illustrating the class usage for image data. Finally, we used this method to understand association between BMI and the weighted loading matrices from the cortical thickness data of human connectome project[4]. We expect this research can be extended for higher dimensional data.

## 2 Non-negative matrix factorization

The NMF method reduces data dimensionality by imposing non-negativity constraints. These constraints lead to a part-based representation because they allow only additive combinations of the original data. The decomposition finds two new matrices **W** and **H** shown in Eq. 1. Each column of **W** contains a basis vector while each column of **H** has the weights to approximate the corresponding column in **X** using the basis vector from the **W** matrix.

The method starts with a user-defined initialization. The initialized matrices are iteratively updated to minimize the Frobenius norm objective function in Eq. 1. In our standard implementation of the projective orthonormal NMF[19] the objective function is minimized subject to the non-negativity constraints on **W** and **H**, the projective constraint in Eq. 2, and the orthonormal constraint in Eq. 3. The projection matrix is set as **P** = **WW**^*T*^ from the constraint **H** = **W**^*T*^ **X** with the objective of approximating **X** as **PX**.

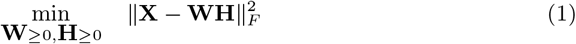

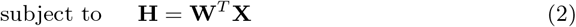

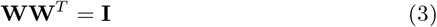

With an appropriate scaling coefficient to the gradient of the cost function, we have the multiplicative update as follows[19]:

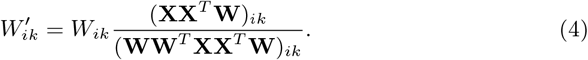

We have two different versions of the multiplicative update depending on the dimension of the input matrix **X**.

if **XX** := **XX**^*T*^ has a small size,

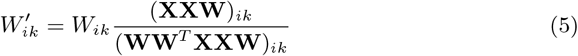

otherwise, **XX** is separated for efficient memory use.

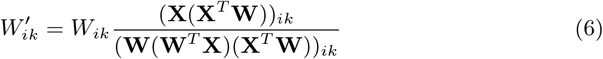

## 3 NMF class design

The interface to the NMF methods was provided via the abstract class defined in itkNMFBase.h [10]. The base class has the common data containers and parameters associated with the decomposition method, such as the input matrix, the decomposed matrices, the number of basis decomposition classes, and the default initialization. The interface for the derived classes via virtual function has been revised for this package. For a specific instantiation of the framework, an implementation of the decomposition by the class named itkEuclideanNMF.h is provided for the Euclidean norm based objective function defined in the previous section.

The decomposition method requires the implementation of a convergence testing that terminates the iterations when no significant changes occur in the decomposed **W** and **H** matrices. The definition of significance is difficult and can change from application to application. In our case, we compute the Frobenius norm of the change in **W** matrix normalized by the norm of **W** matrix itself shown as the variable diffW in the Appendix. The relative norm should be less than the tolerance value for convergence. The iterations also terminate if the method does not converge within a user defined maximum number of iterations.

In this implementation The matrix **W** is initialized by the nonnegative double singular value decomposition (NNDSVD) or its variations[1]. NNDSVD starts either by the partial SVD that computes the largest *K* eigenvalues and their eigenvectors or by its randomized version[5]. Then NNDSVD chooses the larger magnitude of positive or negative eigenvectors.

### 3.1 Modification from ITK NMF Class

The class itkNMFBase now has the additional method called “SetInput” to remove zeros rows and check the negative values from the data matrix. The cost of the Euclidean norm (Frobenius norm) of the factorization error was implemented in the itkEuclideanNMF class that was modified from the itkKullbackLeiblerNMF class that implemented the corresponding cost of the Kullback Leiber divergence between the original matrix and the product of the two factorization matrices because The matrices can be thought as the probability distributions after normalization. Initialization of the factor (**W** matrix) was moved from the itkNMFBase class to the derived class itkEuclideanNMF because the initialization requires the member variables of the derived class, ‘initMeth’ that is not a member of the base class. In addition, many functions have been added to set up the member variables of the derived class. The following methods of TestConvergence(), NMFUpdate() and WriteMLE() were merged into Compute() because they are not complex.

### 3.2 Data Flow

There are two possible ways of inputs and outputs of NMF application. The first one generates the data matrix whose columns correspond to the vectorized input images of each samples. The second one has the data matrix provided directly by the user. The data matrix is loaded by Armadillo. In the same way, the first one outputs the **W** as a number of factor images in either NIFTI or CIFTI format. The second one generates the factor matrices **W** and **H** as ‘csv’ files. For the first case, ‘preprocess image.cxx’ or ‘preprocess surf image.cxx’ reads the NIFTI or CIFTI images to analyze, convert them into the data matrix, then the factor matrices are generated by Armadillo. Similarly, ‘write image.cxx’ or ‘write surf image.cxx’ reads the columns of the **W** matrix and writes them into the image output files in NIFTI or CIFTI format. Figure 1 shows the overall algorithm of the proposed package.

**Figure 1:**
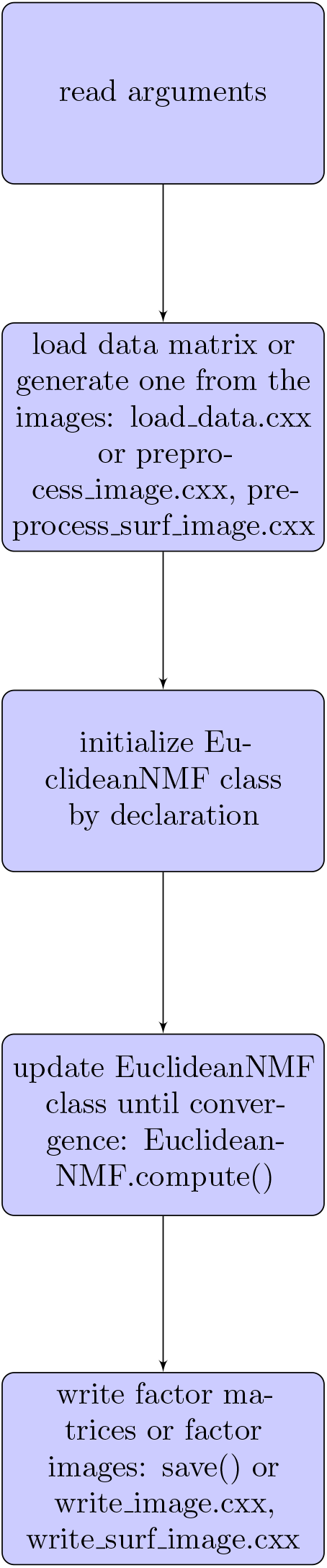
Overall work flow of the software

### 3.3 Usage

The minted box below shows the interface of the NMF decomposition. This example also works as a unit test in the toolkit. The code was successfully compiled with GCC 7.4.0, Armadillo 9.600.6, ITK library 5.0.1, and executed on Ubuntu 18.04.3 LTS. After declaring the itkEulideanNMF class ‘nmf’, the input matrix is set up by the class method ‘SetInput’. Then the class member variables are set up such as the number of classes(factors), the output directory, and the initialization method. The initialization of the class is followed. After the computing the factor matrices in the method ‘Compute()’, its output matrices are read from the class and saved into the csv files.

~~~
**#include** *<*iostream *>*
**#include** *<*armadillo *>*
**#include** “itkEuclideanNMF . h”
**int** main (**int** argc, **char** ∗ argv [])
{
                       **typedef** Mat*<***double***>* InMatrixType, OutMatrixType ;
                       *// Declare the input matrix*
                       **unsigned int** nrows = 4 ;
                       **unsigned int** ncols = 6 ;
                       InMatrixType mat(nrows, ncols) ;
                       *// I n i t i a l i z e the input matrix*
                       mat (0, 0) =1; mat (0, 1) =1; mat (0, 2) =1; mat (0, 3) =2; mat (0, 4) =2; mat (0, 5) =2;
                       mat (1, 0) =4; mat (1, 1) =4; mat (1, 2) =4; mat (1, 3) =3; mat (1, 4) =3; mat (1, 5) =3;
                       mat (2, 0) =4; mat (2, 1) =4; mat (2, 2) =4; mat (2, 3) =3; mat (2, 4) =3; mat (2, 5) =3;
                       mat (3, 0) =1; mat (3, 1) =1; mat (3, 2) =1; mat (3, 3) =2; mat (3, 4) =2; mat (3, 5) =2;
                       *// Declare the decomposition filter*
                       i t k : : itkEuclideanNMF nmf ;
                       *// Set the input matrix*
                       nmf . Set Input (mat) ;
                       *// Set the number of classes in the decomposition*
                       nmf . SetNumber Of Classes (2) ;
                      *// Initialize the filter*
                      nmf . I n i t i a l i z e () ;
                      *// Set the output d i r e c t o r y*
                      nmf . Setoutputimagedir (“. / output”) ;
                      *// Make the output d i r e c t o r y i f i t does not e x i s t*
                      nmf . Makeoutputimagedir () ;
                      *// Set the initialization method*
                      nmf . Setinit Meth (0) ;
                      *// Set the filter to compute the decomposition*
                      nmf . Compute () ;
                     *// Get the results of decomposition*
                     OutMatrixType& W = nmf . GetWMatrix () ; OutMatrixType& H = nmf . GetHMatrix () ;
                      *// write W, H matrices onto csv f i l e s*
                      s t r i n g outputmatrixname = nmf . Getoutputimagedir () +”H. csv”;
                      H. save (outputmatrixname, c s v a s c i i) ;
                      outputmatrixname = nmf . Getoutputimagedir () +”W. csv”;
                      W. save (outputmatrixname, c s v a s c i i) ;
     **return** 0 ;
}
          The NMF application requires the input data and the number of components K as its input. The optional parameters of NMF can be set as follows:
. /NMF **−**help
− example command l i n e o p t i o n s
Usage :
     . /NMF [OPTION …]
     −i, **−**input FILE Input F i l e
−K, Integer Number of factors
[−o, **−**output DIR] Output filedirectory (**default** : . / output)
           [−− intermedia te DIR] directory **for** intermedia te fil es (**default** :“”)
           [−−initW FILE] Initial Wmatrix File (**default** :“”)
           [−− help] Print help
           [−− maxiter Integer] Maximum number of iterations (**default** : 50000)
           [−− initmethod Integer] Method of initialization,
                                                              0 : NNDSVD (**default**),
                                                              1 : NNDSVDa,
                                                              2 : NNDSVDar,
                                                              3 : NNDSVD **using** random SVD calculation,
                                                              4 : random number
[−− iter 0 Integer] Starting iteration number **for** restart (**default** : 1)
[−− saves tep Integer] every s teps to save the intermed iate results (**default** : 0)
[−− t o l **double**] Tolerance o f convergence (**default** : 1e −5)
[−−dim arg] Dimension o f binary input f i l e,
                                        **−**dim=n rows, ncols (**default** : 1, 1)
[−− ciftiflag boolean] CIFTI Flag, ei ther NIFTI (**false** : **default**) **or** CIFTI (**true**)
[−−mem flag boolean] Memory Flag to use the memory savin g code (**default** : **false**)
[−− input flag boolean] Either data matrix (**false** : **default**) **or** list of input images
~~~

## 4 Numerical Results

This section shows the computing performances of the package in speed, accuracy, and memory requirement against MATLAB. In addition, toy examples of simulated images are provided to demonstrate that the package. Finally, the cortical thickness data are analyzed by the package to show its validity from the human connectome project[4].

### 4.1 Computing Performances

This subsection shows that the proposed software is reliable in terms of speed, accuracy, memory requirement, etc. The test was performed by four Intel Xeon Gold 6226 processors with clock speed at 2.70GHZ under CentOS 6.10. The cortical thickness data was processed in the following way. The Multimodal Surface Matching (MSM) surface registration algorithm [12] was used to perform an initial gentle non-rigid surface registration based on folding patterns. This registration, together with the FNIRT nonlinear volume registration[15] to MNI template, was used to bring an initial version of the data into standard grayordinates space. For example, 32k standard mesh for each hemisphere’s cortical surface at 2 mm average vertex spacing and 2 mm isotropic MNI-space[2] voxels for the subcortical volume data. On the other hand, 164k standard mesh for the cortical surface has 0.9 mm average vertex spacing.

The running hours appear consistent with respect to various number of components for a different number of samples in Figure 2. For 40 components, the running hours increase approximately ten hours per 200 subjects. The used memories are proportional to the numbers of samples in Figure 3. In the same way, the used memories are proportional to the size of images whose resolutions are either high or low in Figure 5. The running hours increase in proportion to the data size from their resolution differences in Figure 4.

**Figure 2:**
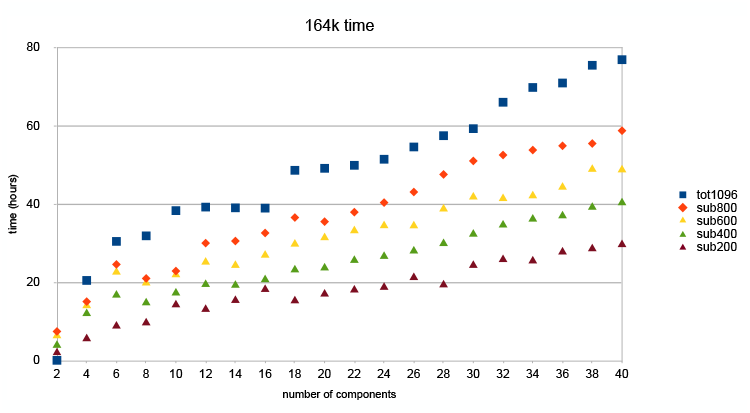
time complexity in cpu hours with various number of components for different number of samples

**Figure 3:**
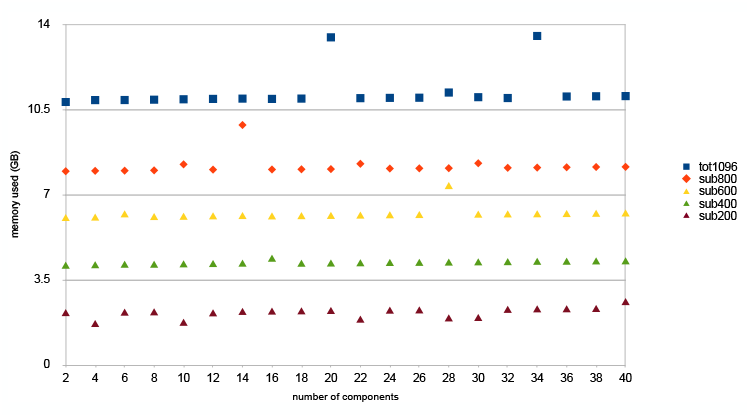
Memory usage with various number of components for different number of samples

**Figure 4:**
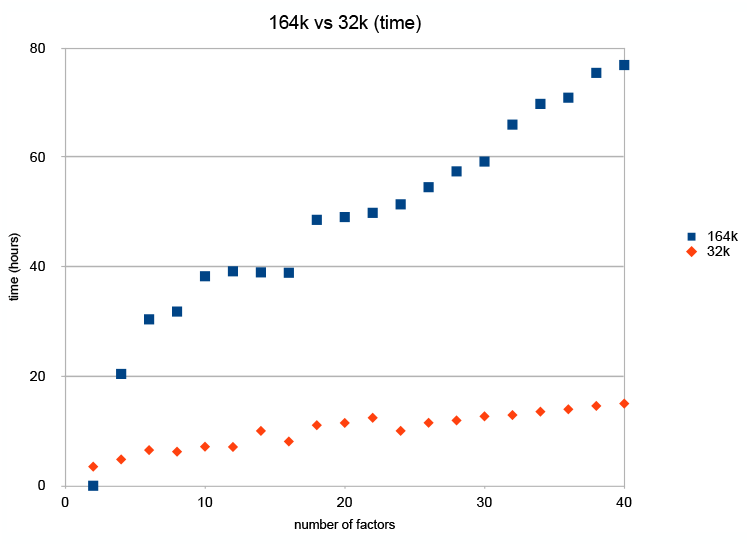
time complexity in cpu hours with various number of components for cortical thickness images of different resolutions, 32k and 164k

**Figure 5:**
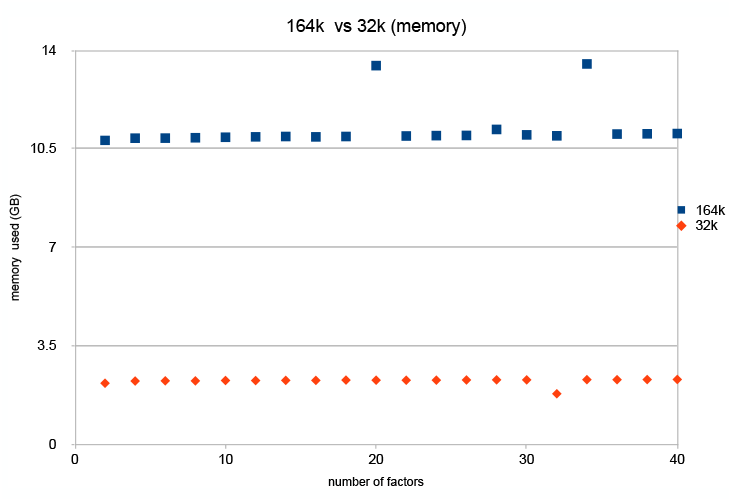
memory usage with various number of components for cortical thickness images of different resolutions, 32k and 164k

The normalized norms of the error are practically negligible in the factor matrix **W** for various numbers of the factors in Figure 6 for both low resolution(32k) images and high resolution(164k) images. The proposed software is slightly slower than MATLAB for a variety of the number of threads for 164k cortical thinkness data that were smoothed with 8mm Gaussian kernel for ten factors in Figure 7. Although the authors of Armadillo claimed its speed excellent, MATLAB was apparently better optimized.

**Figure 6:**
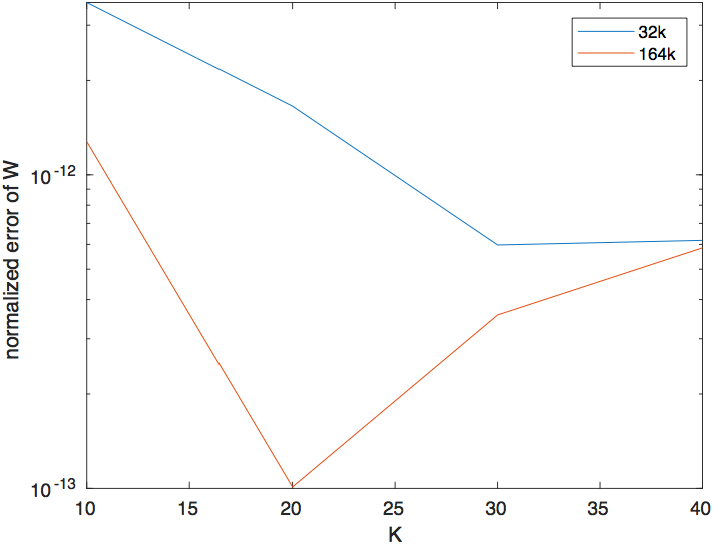
The normalized norms of the errors of the factor matrix **W** for the cortical thickness images with two different resolution, 32k and 164k

**Figure 7:**
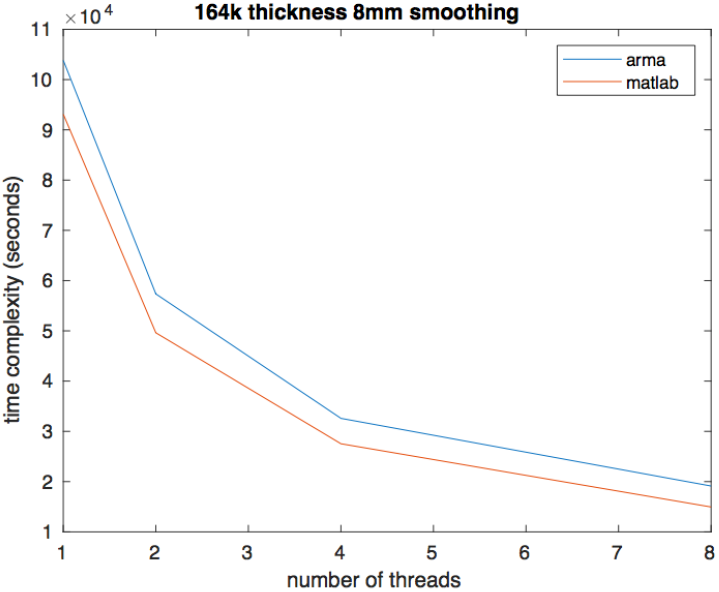
Speed comparison between the proposed package and MATLAB for a various number of threads for 164k cortical thickness data for ten factors.

### 4.2 Examples from Simulated Images

For a validation, we have simulated a set of image data corrupted by noise. The test results were generated using the NMF code compiled with GCC 7.4.0, Armadillo 9.600.6, ITK library 5.0.1, and executed on Ubuntu 18.04.3 LTS. We first generated 2 basis images each of size (256 *×* 256 pixels) with 0 being background (black) and 1 being foreground (white) in Fig. 8 (a) and (b). These are our ground truth images. The pixel intensity of the foreground was scaled to follow the Gaussian distribution with different means varying 100, 125, 155, and 175 for each circles and each group. Then the Gaussian noise with mean 0 and variance 4 was added to the basis images. The examples of the two basis images are shown in Fig. 8 (c) and (e). The underlying pattern is distinguishable in the figure. Then those corrupted images were converted to a vector of length equal to the total number of pixels by raster scanning along each column. 100 copies of the 2 basis vectors were made and arranged in two groups to form an input matrix of dimension 63556 rows and 200 columns (2 groups of 100 images). The recovered basis images from NMF match the original image bases in Fig. 8 (e) and (f).

**Figure 8:**
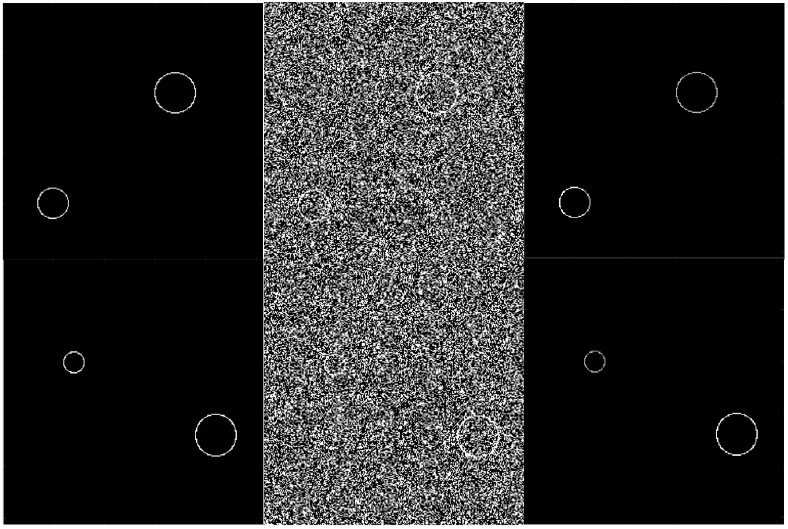
Recovery of noisy image bases from nonnegative factorization. Each columns represent the ground truths, noisy bases, and recovered bases, respectively. Two rows represent two groups of bases.

### 4.3 Application to HCP data

The experimental data were drawn from the human connectome project (HCP), which is a collaboration between Washington University and University of Minnesota. The population-based sample included young adults who volunteered to participate in the studies charting the neural pathways that underlie brain function and behavior, including high-quality neuroimaging data in 1206 healthy young adults. Of these, 110 subjects were excluded because their cortical thickness images did not exist. The resulting sample, which was used for the structural neuroimaging analysis, included 1096 young adults (596 females) ages 22-37 (mean, 28.79 years; SD, 3.69) of mixed race (822 white; 160 black; 114 other) was used.

The image acquisition method is presented in [4]. After acquisition, registration and smoothing operation was applied as described in the subsection 4.1. Then the data matrix was formed by vectorizing the three dimensional voxels of each subject and concatenating them columnwise for every subjects. Then the data matrix was factorized as 16 components by NMF(Figure 11). The number of the components, intrinsic data dimension, were chosen either by finding the inflection point of the decrease of the reconstruction errors(Figure. 9) or by finding the local maximum of the reproducible decompositions from the split half analysis(Figure 10).

**Figure 9:**
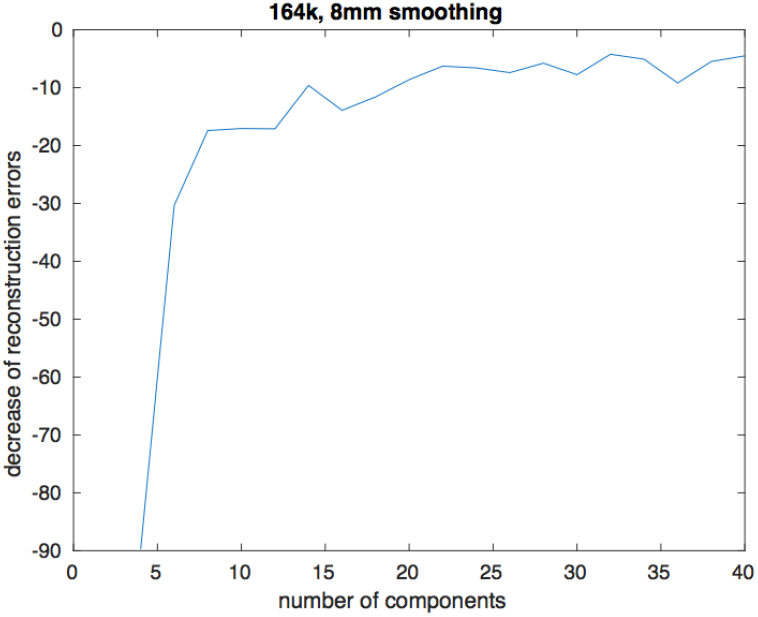
Decrease of the reconstruction errors with increasing number of components for smoothed cortical thickness images of high resolution 164k. We note an infection point for 16 components suggesting that the intrinsic data dimension has been reached

**Figure 10:**
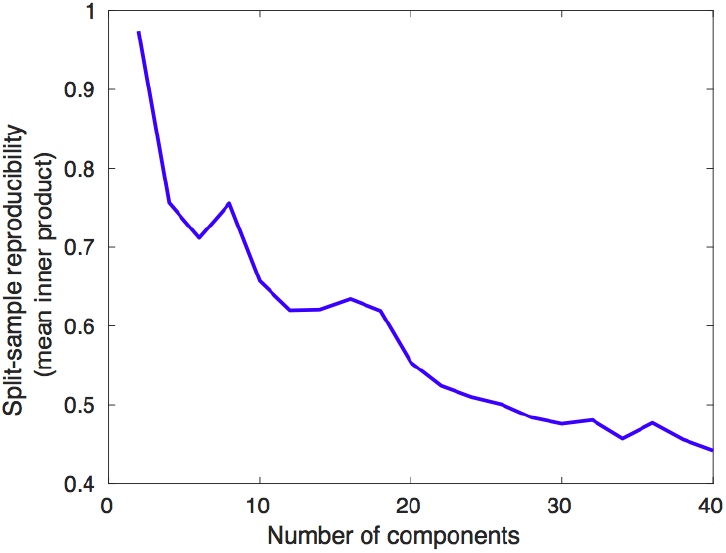
Reproducibility of the decomposition algorithm is plotted as a function of the estimated patterns for the split-half analysis. The decomposition is less stable with increasing number of estimated components, which is expected because the complexity of the problem increases, while the size of the patterns and the modeled effect decreases. The local maxima indicate the number of patterns that can be stably estimated (i.e., 8 and 16 patterns)

**Figure 11:**
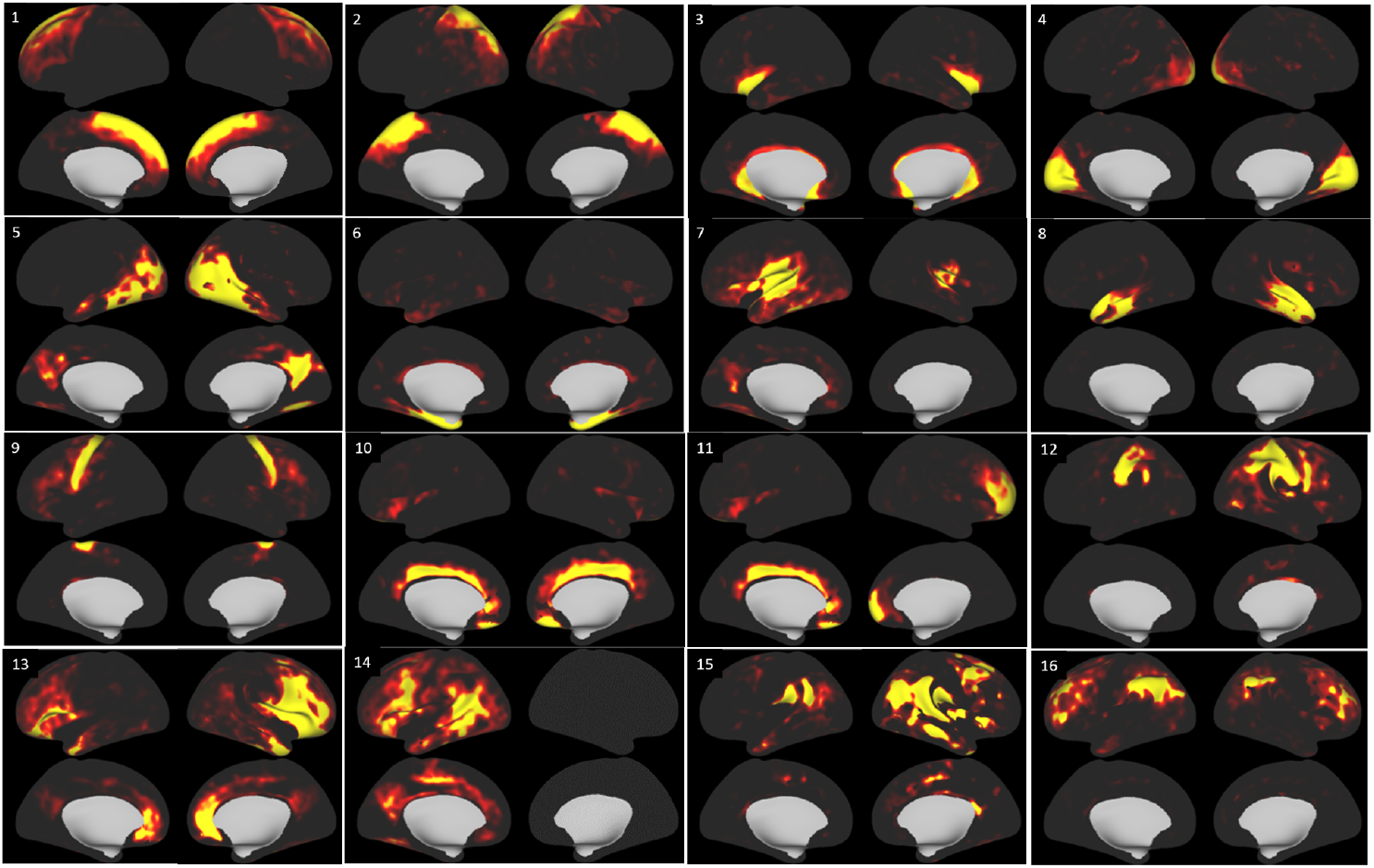
Factor images of the cortical thickness data by the nonnegative matrix factorization. Factors 1-16 are shown in the order of rows. For example, factor 1 and 2 are shown as the left and right images in the first row.

The corresponding loading coefficient matrices were normalized and called the weighted thickness data. Finally, the regression analysis was performed of the weighted thickness data against epidemiological independent variables such as age, gender, and categorized body mass indices such as underweight, normal, overweight, and obese. ‘Underweight’ is defined when the body mass index (BMI) is less than 18.5; ‘normal’ when 18.5 ≤ BMI *<* 25 holds; ‘overweight’ when 25 ≤ BMI *<* 30 holds; ‘obese’ when BMI ≥ 30 holds.

The regression analysis gave all the components significant with respect to age except the sixth factor and four significant factors against gender. There were four significant factors with respect to underweight. Especially, factor 11 matched the dorsolateral prefrontal cortex from [18]. With respect to the product term overweight*gender was significant the factor 4 that corresponds to medial prefrontal cortex in [18, 3]. The same factor 4 was significant for both product terms normal*gender and overweight*gender in Figure 11. The associated false discovery rate (FDR) adjusted pvalues are shown in Table 1.

**Table 1:**
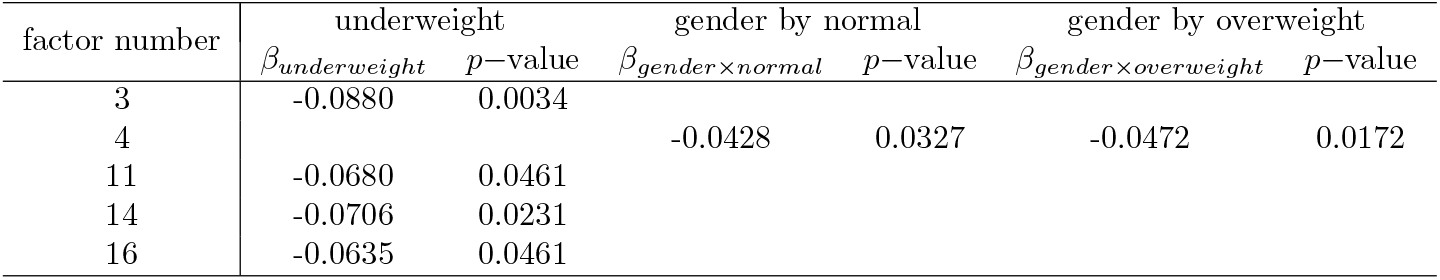
Regression analysis.

## 5 Discussion

We introduced an open-source C++ package for performing NMF based on ITK and Armadillo, a MATLAB style C++ based math library. Armadillo facilitates the computations in linear algebra by calling functions without any need to implement functions in C++. In addition, This framework supports the read and write interface to images specific to neuroscience such as NIFTI and CIFTI. We showed that the package has accuracy matching MATLAB and its speed close to that of MATLAB. We demonstrated that the package can be used to analyze neuroimaging data from the human connectome project. Specifically, we used the package to find a set of structural patterns(factor matrices) that are similar across individuals. We validated this factorization method by associating their weighted loading matrices with body mass indices (BMI) of individuals.

In spite of the above merits, the package does not support many variations of NMF that were implemented in NMFLibrary and NMF toolbox. For example, it support only the multiplicative update and does not cover the alternative least squares method like NMF Library. So there are remaining work to add more functionality to this package in the future.

## Data Availability

All data produced are available online at human connectome db webpage.

https://balsa.wustl.edu/

## Acknowledgments

The authors appreciate Tim Coalson’s kind help to understand and use the Cifti library.

## 6 Appendix

~~~
An Armadillo implementation is compared against a MATLAB implementation to show its similar syntax below. Matrix operations and norm functions from Armadillo match those of MATLAB. Hence Aramdillo has a concise and simple implementation like MATLAB. A MATLAB version is given here.
XX = X ∗ X’;
W = W . ∗ (XX∗W) . / (W∗ (W’ ∗XX∗W)) ;
W(W*<*1e −16)=1e −16;
W = W . / norm (W) ;
  diff W = norm (W old−W, ‘f r o’) / norm (W old, ‘f r o’) ;
       An Armadillo version is given below.
mat XX = X∗ tr a n s (X) ;
W = W % (XX ∗ W) / (W ∗ (tr a n s (W) ∗ (XX ∗ W))) ;
uvec idW = f i n d (W *<* 1e − 16);
W. elem (idW) = 1e −16∗ ones (arma : : s i z e (idW)) ;
W = W / norm (W, 2) ;
diff W = norm (W old−W,”f r o”) / norm (W old,”f r o”) ;
~~~

